# The accuracy of large language models in labelling neurosurgical ‘case-control studies’ and risk of bias assessment: protocol for a study of interrater agreement with human reviewers

**DOI:** 10.1101/2024.08.11.24311830

**Authors:** Joanne Igoli, Temidayo Osunronbi, Olatomiwa Olukoya, Jeremiah Oluwatomi Itodo Daniel, Hillary Alemenzohu, Alieu Kanu, Alex Mwangi Kihunyu, Ebuka Okeleke, Henry Oyoyo, Oluwatobi Shekoni, Damilola Jesuyajolu, Andrew F Alalade

**Author notes:** **Please address all correspondence to:** Dr Joanne Igoli, Deanery of Clinical Sciences, The University of Edinburgh, Edinburgh, United Kingdom. EH16 4SB. Joint-first authors: contributed equally (Joanne Igoli, Temidayo Osunronbi).

## Abstract

**Introduction:** Accurate identification of study designs and risk of bias (RoB) assessment is crucial for evidence synthesis in research. However, mislabelling of case-control studies (CCS) is prevalent, leading to a downgraded quality of evidence. Large Language Models (LLMs), a form of artificial intelligence, have shown impressive performance in various medical tasks. Still, their utility and application in categorising study designs and assessing RoB needs to be further explored. This study will evaluate the performance of four publicly available LLMs (ChatGPT-3.5, ChatGPT-4, Claude 3 Sonnet, Claude 3 Opus) in accurately identifying CCS designs from the neurosurgical literature. Secondly, we will assess the human-LLM interrater agreement for RoB assessment of true CCS.

**Methods:** We identified thirty-four top-ranking neurosurgical-focused journals and searched them on PubMed/MEDLINE for manuscripts reported as CCS in the title/abstract. Human reviewers will independently assess study designs and RoB using the Newcastle-Ottawa Scale. The methods sections/full-text articles will be provided to LLMs to determine study designs and assess RoB. Cohen’s kappa will be used to evaluate human-human, human-LLM and LLM-LLM interrater agreement. Logistic regression will be used to assess study characteristics affecting performance. A *p*-value < 0.05 at a 95% confidence interval will be considered statistically significant.

**Conclusion:** If the human-LLM agreement is high, LLMs could become valuable teaching and quality assurance tools for critical appraisal in neurosurgery and other medical fields. This study will contribute to validating LLMs for specialised scientific tasks in evidence synthesis. This could lead to reduced review costs, faster completion, standardisation, and minimal errors in evidence synthesis.

## Introduction

Observational studies, including cross-sectional, cohort, and case-control studies, are ideal for neurosurgery research when placebo or no-treatment groups are risky or ethically challenging or when randomised controlled trials are impractical due to logistical complexities or inadequacy for addressing clinical questions [1].

Cross-sectional studies concurrently evaluate exposure and outcome status at a single time point without longitudinal follow-up [2]. Cohort studies divide participants based on exposures or treatments and follow them over a period, either prospectively or retrospectively, to compare outcomes between the groups [2]. Case-control studies (CCS) compare individuals with (case) and without (control) a particular outcome, retrospectively examining differences in exposure risk factors [2].

Unlike other observational studies, CCS is best suited for investigating rare outcomes or those with long latency periods, leading to its increasing use in neurosurgery [3]. However, they have limitations such as recall bias and the inability to determine incidence and absolute risk or establish temporality [1–3]. Previous research indicates a significant prevalence of misclassified ‘CCS’ in neurosurgery literature, ranging from 41% to 63% [1–3]. Mislabelling of CCS is not unique to neurosurgery, with mislabelling rates reaching as high as 30% to 97% in other fields [4–6]. Cohort studies are most frequently mislabelled as CCS, leading to a downgrading of evidence quality since cohort studies represent the highest level of evidence among observational studies [1–3].

Moreover, mislabelled CCS often report odds ratios instead of relative risks, leading to distorted effect size measurements, particularly in systematic reviews and meta-analyses [3]. Hence, accurate labelling of study designs is crucial for stakeholders, including readers, authors, and editors. In addition, assessing the risk of bias (RoB) is critical to systematic reviews. This process involves reviewing and understanding each eligible study, which relies on a solid grasp of study methods and RoB assessment tools. However, RoB assessment is labour-intensive and prone to human error, which may introduce biases in the conclusions of an evidence synthesis [7].

The recent upsurge in excitement about artificial intelligence (AI) has increased its impact on every aspect of healthcare [8]. Large Language Models (LLMs), a subset of AI, are trained on extensive amounts of text data to understand, generate, and process human-like language for various natural language processing tasks [8, 9]. Many healthcare professionals have begun to use LLMs such as ChatGPT and Claude as advanced search tools for complex medical information. These models exhibit emergent properties resembling human-level intelligence and have demonstrated impressive performance on various medical speciality exams, including neurosurgery [10, 11], and have even succeeded in challenging tests like the United States Medical Licensing Examination [9]. Additionally, some machine learning systems, such as the RobotReviewer, have shown high accuracy in evaluating the risk of bias in clinical trials [12]. However, the potential of LLMs, an advanced AI tool, in categorising study designs and assessing RoB in neurosurgery research still needs to be explored. Leveraging LLMs in these tasks may lead to reduced review costs, faster completion times, and decreased errors in the assessment process.

This study aims to evaluate the performance of four publicly available LLMs (ChatGPT-3.5 [OpenAI/Microsoft], ChatGPT-4 [OpenAI/Microsoft], Claude 3 Sonnet [Anthropic], and Claude 3 Opus [Anthropic]) in accurately identifying the design of ‘case-control studies’ in the neurosurgical literature. It also seeks to identify predictive study characteristics that affect LLM performance. Additionally, we will evaluate human-LLM agreement in overall and domain-level risk of bias (RoB) assessment using the Newcastle-Ottawa Scale for CCS [13].

## Materials and Methods

### Search strategy

There are no official lists of all neurosurgical journals in the literature, considering the ongoing introduction of new journals. We conducted an online Google search using the phrase ‘top neurosurgery journals’ to compile a speciality list of journals for neurosurgeons. This search yielded a Google Scholar [14] and Welch Medical Library [15] list of neurosurgical journals, which we complemented with additional journals from a previously published article [3]. We excluded the journals that have stopped publications and those with a nursing theme. Our search strategy featured 34 PubMed-indexed journals (**Appendix 1**).

A PubMed/MEDLINE search was performed for all the articles in these thirty-four indexed journals from database inception to 8 June 2024, using the search terms ‘case-control’, ‘case control’, ‘case controlled’, or ‘case-controlled’ in the title or abstract.

### The human reviewers

The assessment team will include a consultant/attending neurosurgeon (AFA), two neurosurgical trainees/residents with training in critical appraisal/ postgraduate certificate in health research and statistics/ Masters of Neurosurgery by Research (TO, OO), and nine medical students/ clinicians who will be trained on critical appraisal prior to commencing this study.

### Eligibility criteria

Only original research articles reported as ‘case-control’ in the titles or abstracts will be included. Reviews, commentaries, letters, genetic studies, animal studies, and cost-effectiveness studies will be excluded. Similarly, articles will be excluded if they lack the term ‘case-control’/ ‘case control’/ ‘case controlled’/ ‘case-controlled’ in their abstract/title or if this term was used in reference to another study. Studies with ambiguous study design labels in their abstract/ title and/or those that use multiple study designs will be excluded (for example: ‘cross-sectional case-control study’, ‘case-control cohort study’, ‘systematic review/ meta-analysis and case-control study’). In addition, articles that are neurology-focused instead of neurosurgery-focused will be excluded.

The titles/abstracts will be screened independently by pairs of authors using the Rayyan software, with a third author (TO) resolving any discrepancies.

### Data extraction

Data extraction from the eligible full texts will be performed by a pair of authors (TO and other authors), with a third author (OO) resolving any discrepancies. The following data will be extracted based on previous related publications [1, 3]:

- Journal name (The journal results will be presented anonymously in the resulting publication).
- Year of publication (<2008, 2008 - 2019, >2019). The STROBE statement was published in 2007, and the last publication on the mislabelling of case-control studies in neurosurgery was published in 2019 [2, 3]. This forms the rationale for the year categories.
- Topic (spine, trauma, vascular, functional/epilepsy, neuro-oncology, paediatrics, skull base, pituitary, hydrocephalus and other).
- Country of origin (based on where the study took place; the first author’s country will be used when the study location is not specified). Countries will be grouped by the number of case-control studies published (Group A: countries with >10 case-control studies; Group B: countries with 5 to 10 case-control studies; Group C: countries with <5 case-control studies). The countries will also be grouped by continents (Africa, Antarctica, Asia, Australia, Europe, North America, and South America).
- Presence or acknowledgement of a case-control expert in the study (such as a statistician, epidemiologist, or one with a master’s degree or equivalent in public health)
- Study design characteristics:
  ○ Aim of study (a): Outcome assessment
  ○ Aim of study (b): Risk factors assessment
  ○ Used logistic regression analysis.
  ○ Reported odds ratio (OR)
  ○ Used survival analysis/Kaplan-Meier curves.
- Terminology of the study:
  ○ The word ‘cohort’ was used in the methods, results, or discussion sections.
  ○ The word ‘outcome’ was used in the results section.
  ○ The word ‘prospective’ or ‘prospectively’ was used in the methods section.
  ○ The word ‘retrospective’ or ‘retrospectively’ was used in the methods section.

### Assessment of study design and risk of bias by human reviewers

The assessment of the study design of the eligible full text articles will be performed by a pair of authors (TO and other authors), with a third author (OO) resolving any discrepancies. The human assessors will classify the studies as ‘true case-control studies’ or ‘non-case-control studies’. A study will be deemed a true case-control study if it comprises three fundamental elements [1]: 1) compares a group of patients with a disease or who have experienced an event with a control group lacking the disease or event; 2) a retrospective evaluation from the time point of a known outcome is made; and 3) focuses on identifying risk factors/associations/causality of the disease or event. The ‘non-case-control studies’ design will be specified as prospective cohort studies, retrospective cohort studies, cross-sectional studies, case series, case reports, randomised clinical trials, and other.

The Newcastle-Ottawa Scale (NOS) will be used to evaluate the risk of bias (RoB) in the true case-control studies [13]. The true case-control studies will be divided into five groups, and the RoB assessment will be performed by a pair of authors, with a third author (TO or OO) adjudicating any discrepancies. Studies with NOS scores of 0-3, 4-5, 6-7, and 8-9 will be considered unsatisfactory, satisfactory, good, and very good quality, respectively.

### Assessment of study design and risk of bias assessment by LLMs

For each eligible article obtained from the abstract/title screening, the methods section will be copied and imputed separately into each LLM (ChatGPT-3.5 [OpenAI/Microsoft], ChatGPT-4 [OpenAI/Microsoft], Claude 3 Sonnet [Anthropic], and Claude 3 Opus [Anthropic]) and the LLMs will be prompted with this question: ‘Some authors may or may not correctly label their study design. Using the hierarchy of evidence, with a rationale, what is the actual specific study design in the text below?’ To facilitate the assessment of the LLM-LLM intrarater agreement, we will obtain LLM assessments in duplicate, i.e., two different authors (TO and OO) will separately use the LLMs independently for the assessment of study design.

Subsequently, we will evaluate the LLMs RoB assessment for the author-labelled true CCS. The PDF files of the eligible papers will be imputed separately as attachments into each LLM. The LLMs will be prompted with this question: ‘Given that studies with an overall Newcastle-Ottawa scale (NOS) scores of 0-3, 4-5, 6-7, and 8-9 are considered unsatisfactory, satisfactory, good, and very good quality, respectively, provide a domain-level and overall risk of bias assessment for the following study using the Newcastle-Ottawa scale for case-control studies.’ If we are unable to attach the PDF file or the LLM is unable to read the PDF file, we will copy the methods text and the patients/participants characteristics/demographics subsection of the results and impute this into the LLM. To facilitate the assessment of the LLM-LLM intrarater agreement, we will obtain LLM RoB assessments in duplicate — i.e., two authors (TO and OO) will each use the LLMs independently for the RoB assessment.

### Statistical analysis and reportingxs

Statistical analyses will be conducted on IBM SPSS Statistics 27 (Windows).

LLM-LLM and human-human interrater reliability for the study design and RoB assessments will be assessed using Cohen’s kappa (κ) for categorical data. In the event of LLM-LLM (for example, ChatGPT-3.5 - ChatGPT-3.5) discrepancies, we will reduce the duplicate assessments to a single assessment for each study by randomly choosing one of the assessments for each study.

We will calculate the proportion of articles labelled as ‘case-control’ in the title/abstract that are true case-control studies as identified by human reviewers. Furthermore, using the study design determined by human reviewers in this study as a reference, we will calculate the proportion of study design correctly labelled by each LLM. Subsequently, LLM-human inter-rater reliability for the study design and RoB assessments will be assessed using Cohen’s kappa (κ) for categorical data. Kappa values will be interpreted as follows: values ≤ 0 (no agreement), 0.01–0.20 (slight agreement), 0.21–0.40 (fair agreement), 0.41– 0.60 (moderate agreement), 0.61–0.80 (substantial agreement), and 0.81–1.00 (almost perfect agreement) [16].

Simple logistic regression analyses will be conducted to assess the associations between select study characteristics and whether a study was a true case-control (yes/no). These analyses will also be conducted for each LLM to assess the association between the select study characteristics and the accurate labelling of study designs by the LLM (yes/no). A *p*-value < 0.05 at a 95% confidence interval will be considered statistically significant.

## Discussion

To our knowledge, this study will be the first to evaluate interrater agreement between human reviewers and LLMs in labelling study designs and assessing RoB in neurosurgical case-control studies.

If the human-LLM interrater agreement is almost perfect, then LLMs could become valuable tools for teaching and quality assurance in critical appraisal and identifying study designs in neurosurgery and other fields. This study is expected to make a significant early contribution to the research exploring the utilisation and validation of general-purpose LLMs trained on vast internet data for specialised scientific tasks. It is anticipated that this study will mark the beginning of a series focused on employing LLMs in evidence synthesis. The investigation into the application of LLMs, particularly for systematic reviews, is poised to bring about significant changes in how evidence synthesis tasks are conducted, who undertakes them, the speed and cost of completion, and the way primary studies are conducted and reported to enhance comprehensibility for artificial intelligence.

### Limitations

This study will not include some non-neurosurgical-specific journals where the neurosurgical community may choose to publish. Thus, the representativeness of the selected articles as a sample of all neurosurgical case-control studies can be questioned. Based on our exclusion criteria, articles lacking explicit mention of “case control” in title or abstract will be excluded. However, the improper use of the term “case-control” might be more prevalent in these articles and missed in our search. Though unlikely, reverse mislabelling could occur, where true case-control studies may not have been labelled as such and thus missed in our search.

To evaluate LLMs’ ability in RoB assessment, we will provide only the methods and results section or full articles (where possible) to the LLMs. Human reviewers will have access to the entire text and supplementary materials where available, providing them with more information about each study than LLMs. As a result, the human-LLM interrater agreements we estimate are expected to be conservative estimates of what is achievable.

## Data Availability

No datasets were generated or analysed during the current study. All relevant data from this study will be made available upon study completion.

## Declarations

### Authors’ contributions

Joanne Igoli, Temidayo Osunronbi, Olatomiwa Olukoya, Damilola Jesuyajolu, and Andrew F Alalade contributed to the study’s conception and design. The first draft of the paper was written by Temidayo Osunronbi, and all authors commented on the subsequent versions of the manuscript. All authors read and approved the final manuscript.

